# Unveiling the threat: Characterization of *Clostridioides difficile* Infection in the Northwest Region of Buenos Aires between 2019-2023 and Associated Risk Factors redefined through a Meta-Analysis

**DOI:** 10.1101/2024.04.25.24306385

**Authors:** Angela María Barbero, Nicolás Diego Moriconi, Sabina Palma, Josefina Celano, María Gracia Balbi, Lorenzo Sebastián Morro, María Martina Calvo Zarlenga, Jorgelina Suárez, María Guadalupe Martínez, Mónica Graciela Machain, Carlos Gabriel Altamiranda, Gabriel Erbiti, Rodrigo Emanuel Hernández Del Pino, Virginia Pasquinelli

## Abstract

*Clostridioides difficile* stands as the leading cause of hospital acquired enteric infection in developed countries. In Argentina, the epidemiology of *Clostridioides difficile* infection (CDI) is currently poorly characterized. Therefore, we conducted a retrospective case-control study evaluating the prevalence of CDI in 249 stool samples collected between 2019 and 2023 in the Northwest region of Buenos Aires. The presence of *C. difficile* was detected by combining three techniques (EIA, PCR and toxigenic culture) in a diagnostic algorithm. Clinical and demographic data from patients was also analyzed to identify CDI-associated risk factors. 1 in 5 patients presented *C. difficile* as the etiological agent of diarrhea and the 80% of CDI+ cases carried toxigenic strains, with a third of cases acquired in the community. Age ≥69 years, previous use of antibiotics, previous hospitalization and previous episodes of CDI emerged as predisposing factors for CDI in our study cohort. Blood parameters such as an elevated number of leukocytes and platelets, a decreased basophil count, and an increased urea concentration were identified as indicators of CDI. We also carried out a systematic review and a meta-analysis where we contrasted our results with 39 studies selected from different countries around the world. At the global level, the meta-analysis highlighted advanced age, previous consumption of antibiotics and previous hospitalization as CDI risk factors and the leukocyte count as an indicator of CDI. These results emphasize the importance of epidemiological studies and reveal crucial information for healthcare decision-making regarding CDI.

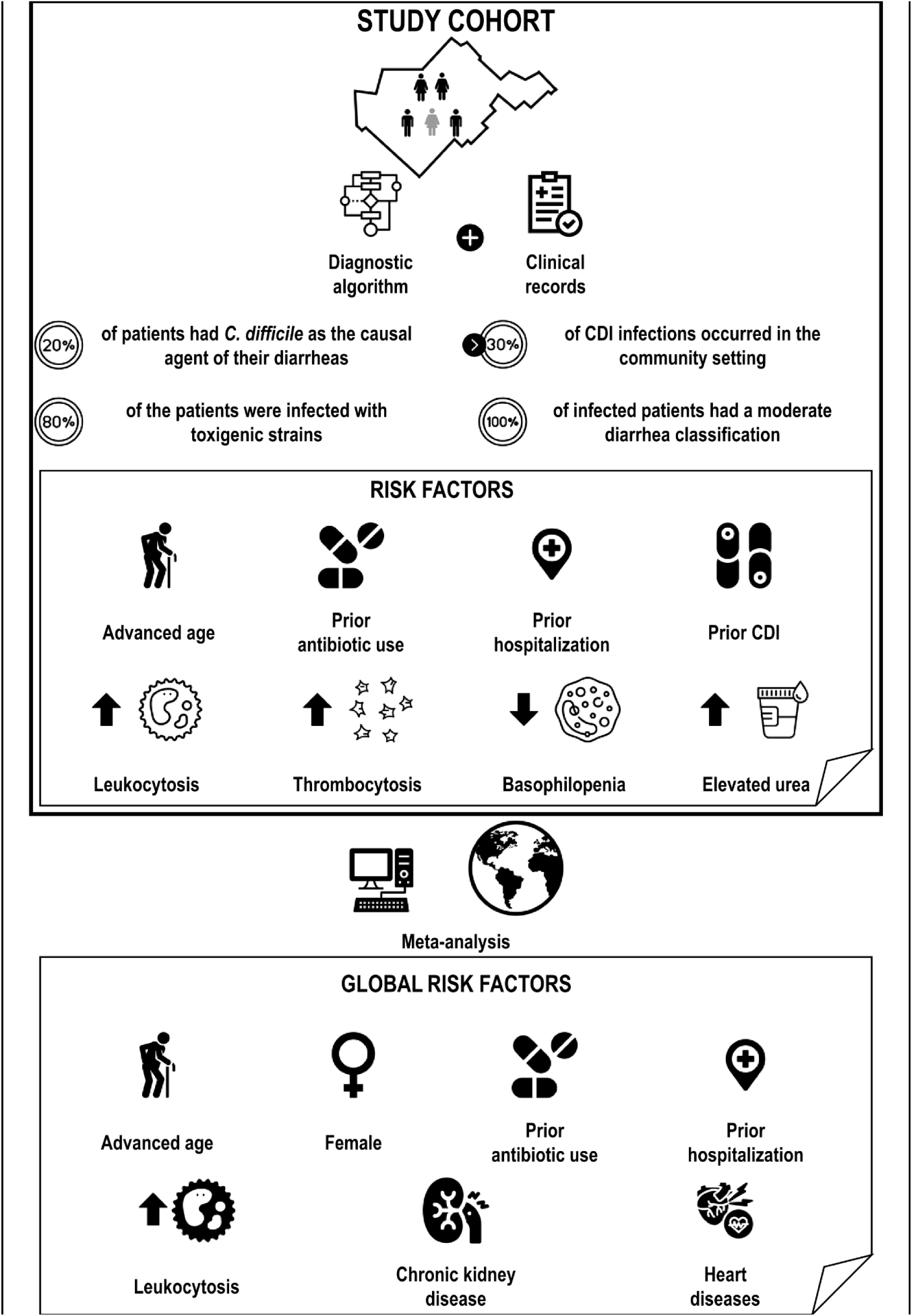

## Introduction

*Clostridioides difficile* infection (CDI) is considered as the most frequent hospital-acquired disease ^1,2^. Since 2019, the Centers for Disease Control (CDC) have identified *C. difficile* as an “urgent threat”, with the immediate need to implement prevention and control actions ^3^. *C. difficile* is a Gram positive, anaerobic and spore-forming bacterium that causes intestinal damage primarily through the production of two main toxins: toxin A (TcdA) and toxin B (TcdB) ^4^. These toxins play a crucial role in the pathogenesis of CDI by damaging the cells lining the gut. CDI can be life-threatening, ranging from mild diarrhea to pseudomembranous colitis and causing nearly 500,000 cases per year in the United States ^5^.

CDI is also a major economic burden to the health care systems, which is closely linked to the high recurrence rates of this infection. Although standard treatments resolve CDI in most cases, up to 35% of CDI-treated patients will experiment a recurrence of the disease with aggravated symptoms ^5^. Common therapy for CDI includes antibiotics such as metronidazole for mild cases and vancomycin or fidaxomicin for moderate to severe cases ^6^. Therapies with the human antibody bezlotoxumab against *C. difficile* toxin B or Fecal Microbiota Transplant (FMT) to restore the balance of the microbiome have been recommended to treat or prevent recurrences ^7,8^.

Regarding global epidemiology, while healthcare-associated CDI has declined in recent years, the incidence of community-acquired CDI is on the rise ^9–14^. The main transmission route for *C. difficile* is through direct Person-to-Person contact by the fecal-oral route ^15^. *C. difficile* spores constitute the main form of resistance and can persist in the environment for long periods ^16,17^. Then, contaminated surfaces, food or water, as well as asymptomatic carriers, are typical sources of community transmission ^18–20^.

Several risk factors are associated with the development of CDI. The use of antimicrobials with emphasis on broad-spectrum antibiotics, has been described as the most significant risk factor for CDI ^21,22^. These antibiotics can disrupt the normal balance of bacteria in the gastrointestinal tract, generating dysbiosis and so allowing *C. difficile* overgrow in the gut. Hospitalization in healthcare facilities and advanced age, which can be related to weakened immune systems and a higher likelihood of residing in healthcare settings, also constitute important risk factors ^22^. The use of stomach acid regulators, such as proton pump inhibitors (PPIs) and H2 blockers, has been associated with an increased risk of CDI ^23^; however, this is still under debate. Previous episodes of CDI increase the risk of recurrence ^24^. Finally, several underlying health conditions that could compromise the immune system could contribute to increased susceptibility or severity of the infection. Comorbidities such as Inflammatory Bowel Disease (IBD) ^25^, Crohn’s disease ^25^, ulcerative colitis ^25^, diabetes ^26^ and chronic kidney disease (CKD) ^27^ can enhance the risk of CDI.

Certain clinical procedures (e.g. chemotherapy treatments and gastrointestinal surgery), malnutrition or enteral nutrition, organ transplantation along with immunosuppressive medications and blood disorders may also elevate the risk of CDI ^28,29^.

Understanding geographical variations in prevalence by studying CDI epidemiology and risk factors is essential for preventing, controlling, and managing the infection. In Latin America, and particularly in Argentina, comprehensive epidemiological data on CDI are limited. It has been reported that patients with diarrhea are not routinely tested for *C. difficile* in developing countries and, when tested, very often only the enzyme immunoassay (EIA) is used ^30–32^. This could lead to an underestimation in the diagnosis and high economic burdens for the health system. Our retrospective study (2019-2023) assesses the prevalence of *C. difficile* among health care centers of the Northwest region of Buenos Aires, Argentina. Furthermore, with the aim to characterize our study cohort, we analyzed the demographic and clinical data of the patients and we conducted a meta-analysis to compare our findings with global reports.

## Methodology

### Human Samples and Participating Institutions

The research was carried out in accordance with the Declaration of Helsinki (2013) and approved by the UNNOBA (Universidad Nacional del Noroeste de la Provincia de Buenos Aires) Ethics Committee (COENOBA).

Fecal samples from hospitalized adult patients with diarrhea were collected after obtaining informed consents and frozen at -20 °C until use. The samples were derived from the following health-care centers: Hospital Interzonal General de Agudos (HIGA) Abraham Félix Piñeyro, Clínica Centro Médico Privada SRL and Clínica IMEC. Samples received between January 1, 2019 and December 31, 2023 from Sanitary Region III of Buenos Aires, Argentina **(Supplementary Fig. 1)** were included in this analysis.

### Patients characterization

The presence of *C. difficile* was determined in the fecal samples using a diagnostic algorithm **(Supplementary Fig. 2)** as recommended by Crobach et al. in 2016 ^33^. Briefly, three tests were used in a retrospective approach: Enzyme immunoassay (EIA, CoproStripTM *C. difficile* GDH + Toxin A + Toxin B (Savyon® Diagnostics Ltd)), Polymerase Chain Reaction from stool samples (PCR, Taq Phire Tissue Direct PCR Master Mix (Thermo Fisher)) and Toxigenic culture (stool culture in CHROMAgar^TM^ *C. difficile* plates + PCR from isolated *C difficile* colonies).

Patients were defined as CDI+ or CDI- and clinical, demographic and blood parameters were evaluated **(Table 1)**.

**Table 1.** Clinical, demographic and blood parameters evaluated in CDI+ and CDI-patients.

| Clinical-demographical parameters | Blood parameters |
| --- | --- |
| Age | Leukocytes (cells/mm<sup>3</sup>) |
| Biological Sex | Neutrophils (cells/mm<sup>3</sup>) |
| Previous hospitalization | Monocytes (cells/mm<sup>3</sup>) |
| Previous consumption of Antibiotics | Lymphocytes (cells/mm<sup>3</sup>) |
| Antibiotics use during hospitalization | Eosinophils (cells/mm<sup>3</sup>) |
| Presence of comorbidities | Basophils (cells/mm<sup>3</sup>) |
| Previous consumption of Proton Pump Inhibitors | Platelets (cells/mm<sup>3</sup>) |
| Previous CDI | Urea (mg/dl) |
| Place of hospitalization | Creatinine (mg/dl) |
| Need for Intensive Care Unit (ICU) | Albumin (g/dl) |
| Origin |  |
| Diarrhea classification |  |
| Shock |  |
| Death |  |

CDI classifications by setting of acquisition and severity were defined according to the Infectious Diseases Society of America (IDSA) criteria ^34^.

### Meta-analysis design

A bibliographic search was carried out using PubMed and Google Scholar databases in addition to the AI SciSpace tool. The reports that fulfilled the definition criteria of cases and controls were selected. The meta-analysis workflow is summarized in **Fig. 4**.

### Statistical Analysis

For clinical, demographic and blood parameters comparisons, parametric t-test or non-parametric Mann-Whitney test for unpaired samples were used. Fisher’s exact test was used to analyze the frequency distribution of qualitative/nominal variables. Data was analyzed using GraphPad Prism 8.0.1 software (San Diego, CA, USA) and p values <0.05 were considered statistically significant.

For the meta-analysis, the “Metafor” package from RStudio (2023.06.1+524 version) was used ^35^. The Odds Ratio (OR, for categorical variables) and the Standardized Mean Difference (SMD, for continuous variables) were calculated and a REML (Random Effect Maximum Likelihood) model was employed. Models with a p value <0.05 were considered as potential predictors of CDI incidence risk.

## Results and Discussion

### CDI prevalence in Northwest Buenos Aires

In Argentina, studies and reports on CDI are scarce and heterogeneous ^36–39^. In order to contribute to a better understanding of the disease, we conducted a retrospective analysis of 249 fecal samples received between 2019-2023 from Sanitary Region III of Buenos Aires.

We determined *C. difficile* as the causal agent of one in five diarrheas (21.29%) **(Fig. 1 a)** and we detected that more than the 80% of the patients were infected with toxigenic strains; indicated by the presence of *C. difficile* Toxin B in the stool samples **(Fig. 1 b)**. This percentage was similar to the worldwide reported prevalence, where CDI is the underlying cause of 15 to 20% of diarrhea associated with the use of antibiotics ^40^.

**Figure 1.**
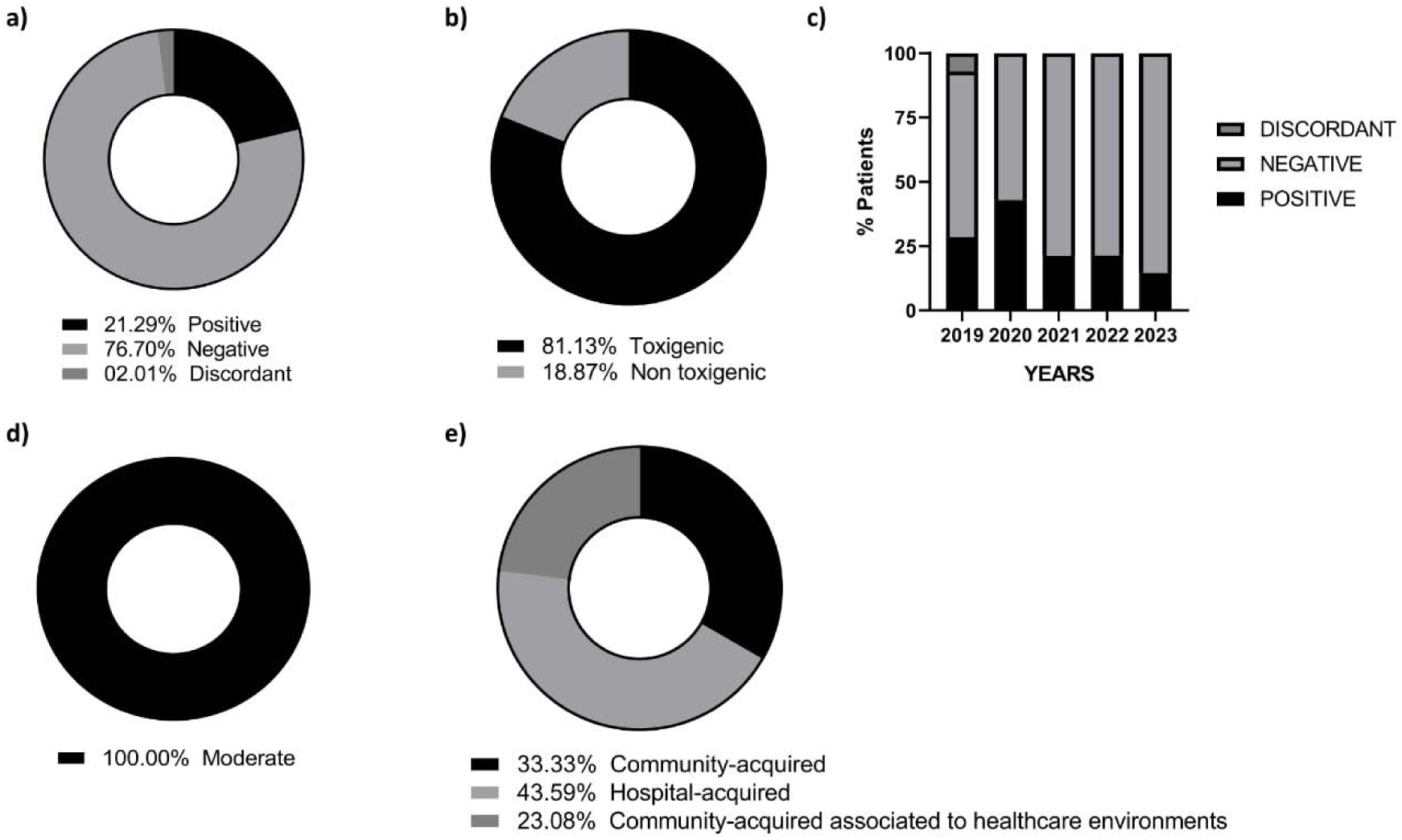
CDI prevalence in Northwestern Buenos Aires. 249 stool samples were evaluated using the diagnostic algorithm. a) Donut graph showing positive, negative and discordant results for the presence of *C. difficile*. b) Donut graph showing the percentage of toxigenic strains (presence of Toxin B) detected in the stool samples. c) CDI+ and CDI-results classified on an annual basis from 2019 to 2023. d) CDI classification. e) CDI classification by setting of acquisition.

When analyzing the cases on an annual basis, the frequency of CDI ranged around 20% from 2019 to 2023 **(Fig. 1 c)**. Interestingly, the highest incidence in our study cohort was observed during 2020, duplicating the percentage of CDI+ cases compared to the rest of the years **(Fig. 1 c)**. In relation to this, the COVID-19 pandemic brought about the preventive use of broad-spectrum antibiotics to avoid bacterial coinfections; which could be associated with the increase in detection ^41–44^. Nevertheless, CDI detection could have been impacted by the decrease in the number of tests, a fact that was corroborated in multiple studies carried out during the early stages of the pandemic ^45–47^. Additionally, there could also be an underestimation of CDI cases ^48,49^ since SARS-CoV-2 frequently causes gastrointestinal symptoms similar to those of C*. difficile* ^50^.

Regarding the severity of the diarrhea, all infected patients had a moderate classification **(Fig. 1 d)**. Importantly, the majority of CDI infections occurred in the community setting (56.41%) (**Fig. 1 e**). 33.33% of the patients presented with community-acquired CDI, 23.08% with community-acquired CDI associated with healthcare environments, and the remaining 43.59% of the patients were hospital-acquired (**Fig. 1 e**). This is particularly notable, since the epidemiology of CDI has changed in the last two decades. The 027 strain, responsible for clinical outbreaks in the early 2000s ^51^, has recently decreased its incidence in part due fluoroquinolones restriction, prevention measures and to improvements in detection tests that have allowed a better characterization of the circulating ribotypes ^52–56^. Additionally, an increase in strains associated with community infection (e.g. 078, 014 in Europe and 106 in the US) has been reported in recent years, with community-acquired cases rising above 40% ^11,57–59^.

### Risk factors

After classifying the patients as CDI+ or CDI-, we determined the risk factors that could be involved in the prevalence of CDI.

As seen in **Fig. 2 a**, we found significant differences in the age of CDI+ and CDI-populations, with the CDI+ patients presenting a higher average age (CDI+: mean = 72 years vs. CDI-: mean = 65 years). By using a ROC curve analysis, we established a cut-off point of 69 years for advanced age as a risk factor in our cohort **(Fig. 2 b)**. Elderly patients have a greater probability of receiving broad-spectrum antibiotics, being hospitalized or staying longer in hospital settings partly due to the presence of comorbidities ^60,61^.

**Figure 2.**
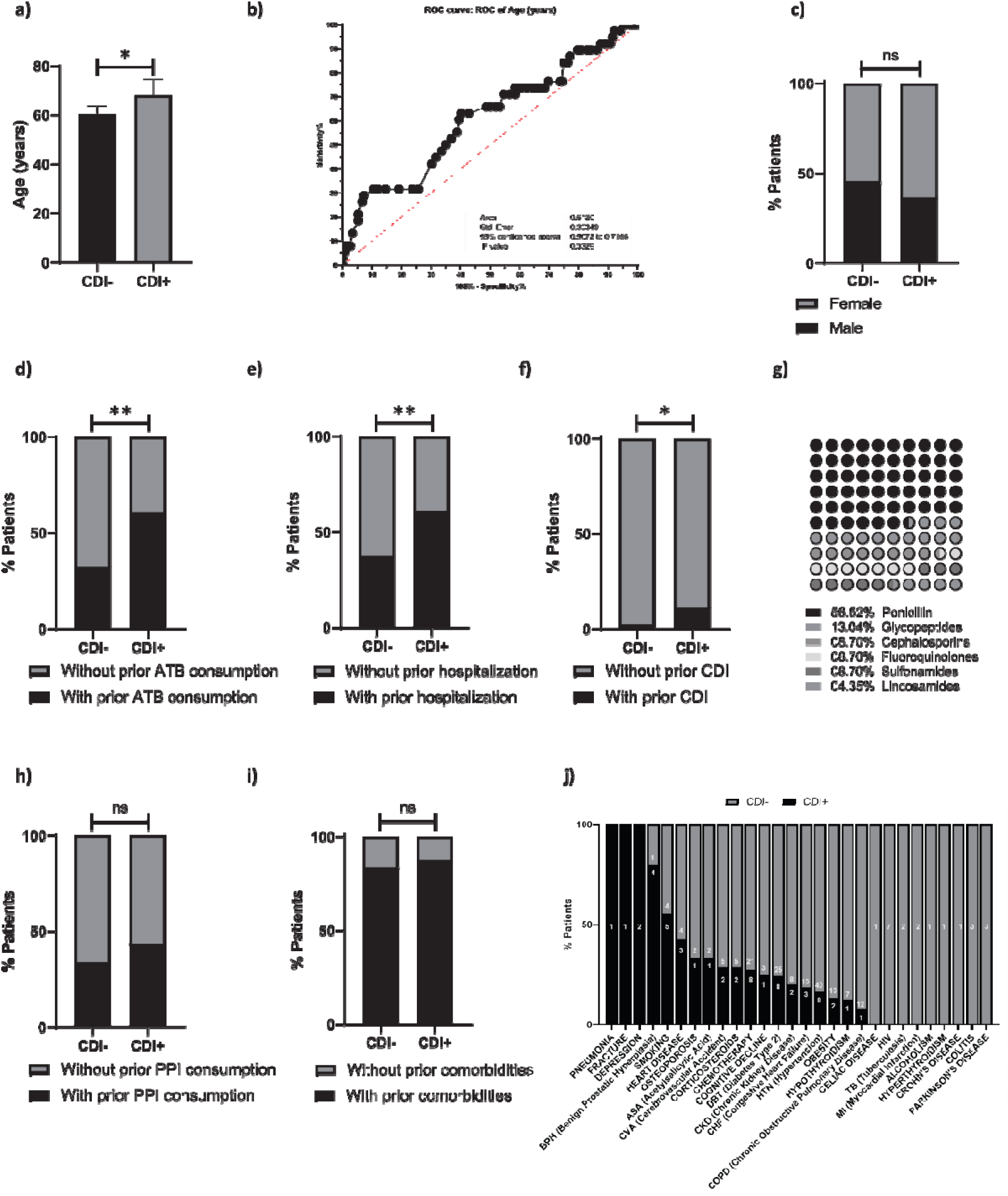
CDI associated risk factors. Evaluation of risk factors associated with CDI. a) Age (years), b) ROC curve to establish the age cut-off point, c) biological sex, d) prior antibiotics (ATB) consumption, e) prior hospitalization, f) prior CDI, g) antibiotic breakdown by family, h) prior PPI consumption, i) comorbidities, j) breakdown of specific comorbidities (the white number inside the bars represents the number of patients with the comorbidity). a) Mann-Whitney test. c, d, e, f, h, and i) Fisher’s exact test. Stacked bars represent the percentage of patients for each parameter. ns= non-significant; *, p

With age, the immune system decreases its functions and also older individuals develop inflammageing, which could impact in the resolution of the infection; a fact that was observed for neutrophils ^62^ and serum IgG against *C. difficile* toxins ^63–65^.

Regarding biological sex, no differences were found for the proportion of males and females between both patient populations **(Fig. 2 c)**.

Consumption of antibiotics in the 3 months prior to the diagnosis of CDI, as well as previous hospitalizations and infections with *C. difficile*, could be considered risk factors that predisposes to CDI in our study cohort **(Fig. 2 d, e and f)**. Hospitals and healthcare facilities are common environments for *C. difficile* transmission since they can be easily colonized by spores that persist on surfaces for months ^66,67^. Previous consumption of antibiotics is directly related to the dysbiosis of the microbiota that enables the colonization of *C. difficile* and is typically consider as the main risk factor for CDI ^68,69^. Some antibiotics, such as clindamycin, broad-spectrum penicillins, cephalosporins and fluoroquinolones alter the microbiota to a greater extent than others ^68,70,71^. When analyzing the families of antibiotics consumed by CDI+ patients, we observed that 56.52% had taken some kind of penicillin prior to diagnosis **(Fig. 2 g)**.

On the other hand, the consumption of proton pump inhibitors (PPI) prior to diagnosis and the presence of comorbidities did not show significant differences between CDI- and CDI+ patients **(Fig. 2 h and i)**. **Fig. 2 j** shows a breakdown of the comorbidities reported in the patients under study. Although they were analyzed individually, no substantial variations were found between the CDI+ and CDI-population for any of them.

No differences were found regarding the variables referring to the evolution of patients during admission to health centers such as *hospitalization in common floor or ICU, need for ICU, shock and death* **(Supplementary Fig. 3)**.

### Blood and serum parameters

We also analyzed the patients’ blood counts and serum parameters that were measured on the day of fecal sample collection.

We observed a significant increase in the number of leukocytes **(Fig. 3 a)** and platelets **(Fig. 3 j)** in patients infected with *C. difficile*. When analyzing the count of lymphocytes, monocytes and neutrophils individually between both patient populations, although an increase was evident, we did not find significant differences **(Fig. 3 b, c and d)**. However, it is important to note that CDI+ patients had increased at least two of these leukocyte populations compared to CDI-patients at the time of diagnosis **(Fig. 3 e, f and g)**. Although some studies have highlighted that an elevated white blood cell (WBC) count is frequently observed in the context of CDI ^72^ ^73^ ^74^, little has been explored in using elevated WBC count as a predictor of this infection. In this regard, we agree with Vargas et al. ^75^ in that the total leukocyte count alone is not a specific indicator for CDI. Previous work evaluating platelet count in CDI episodes reported controversial results, assigning them both a beneficial and detrimental role in relation to clinical symptoms ^76–85^. We have recently shown that platelets bind to *C. difficile* and promote its uptake by human macrophages using macropinocytic pathways ^86^. Therefore, we consider that platelets in CDI could be fundamental for the resolution of the infection and that further studies are needed to unravel their role during CDI.

**Figure 3.**
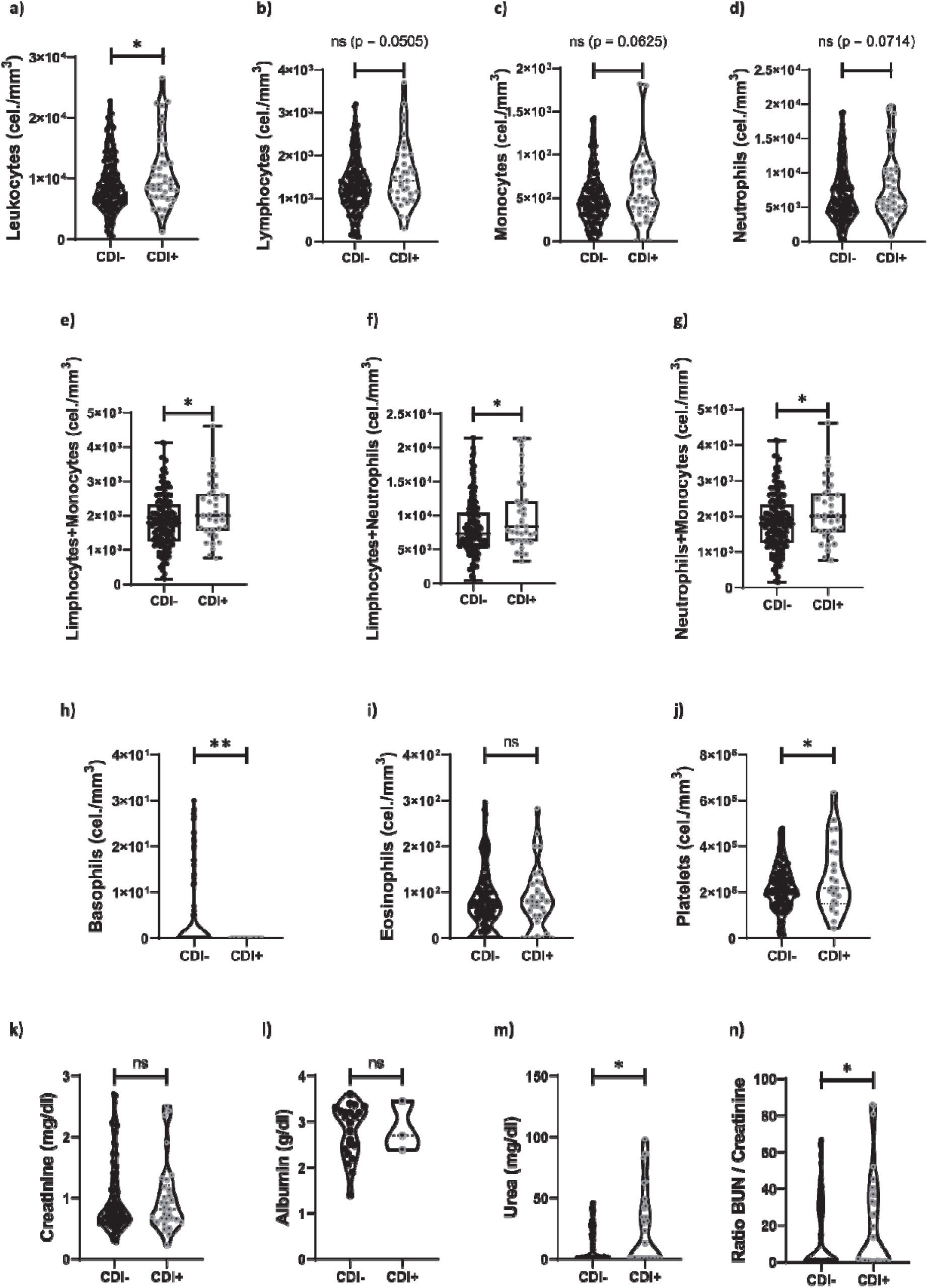
Blood and serum parameters in CDI+ and CDI-patients. Number of a) Leukocytes (cells/mm^3^), b) lymphocytes (cells/mm^3^), c) monocytes (cells/mm^3^), d) neutrophils (cells/mm^3^), e) lymphocytes plus monocytes (cells/mm^3^), f) lymphocytes plus neutrophils (cells/mm^3^), g) neutrophils plus monocytes (cells/mm^3^), h) basophils (cells/mm^3^), i) eosinophils (cells/mm^3^) and j) platelets (cells/mm^3^). Levels of k) creatinine (mg/dl), l) albumin (g/dl) and m) urea (mg/dl). n) BUN (blood urea nitrogen)/creatinine ratio. a-i and k-n) Mann-Whitney test. j) unpaired t test. Violin plots show the distribution of the data. ns= non-significant; *, p<0.05; **, p<0.01.

**Figure 4.**
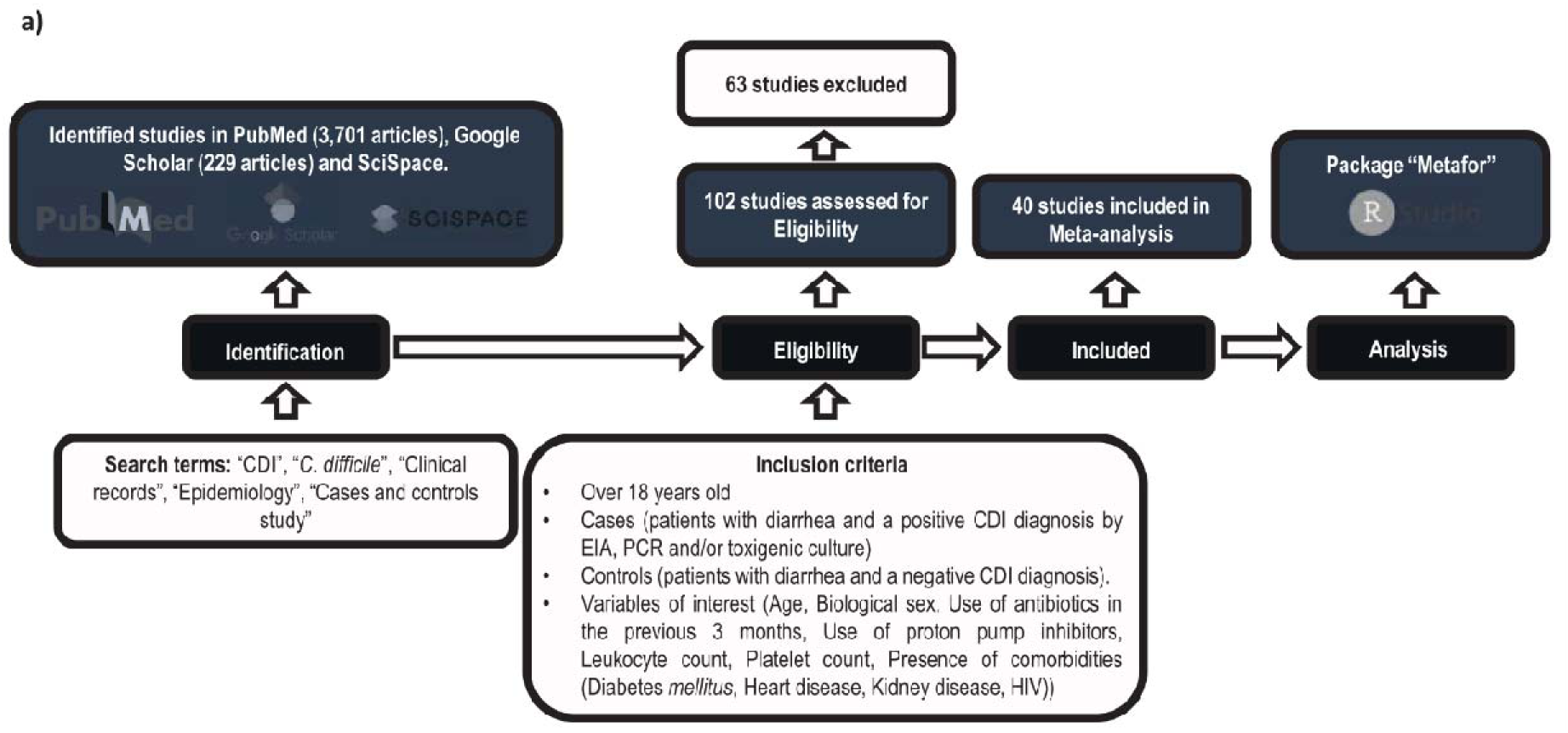
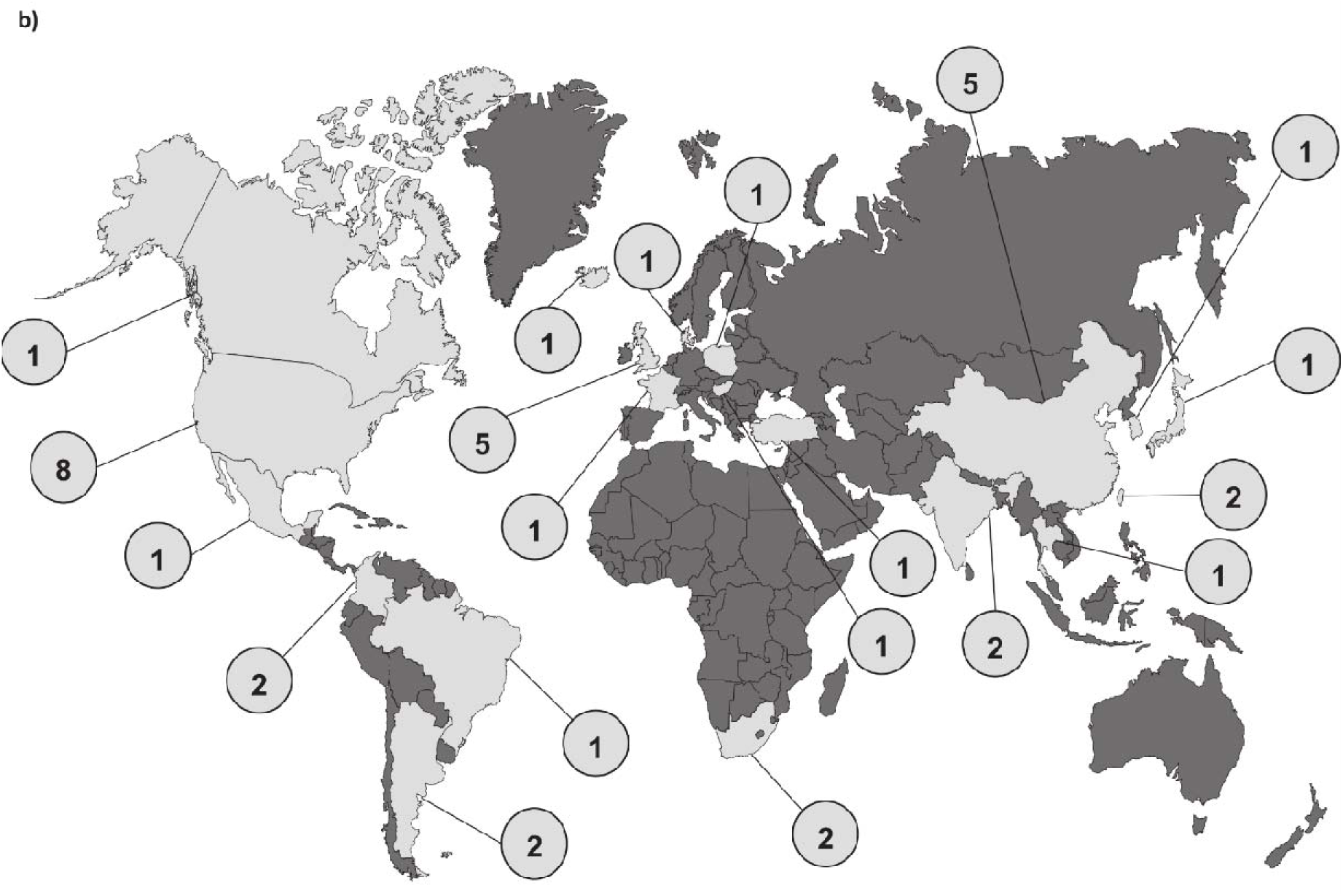
Flow chart of the studies and countries screened and included in the Meta-analysis. a) The obtained results regarding clinical and demographic characteristics of the CDI+ and CDI-patients in our cohort were contrasted against findings of other studies through a meta-analysis by RStudio package “Metafor”. After initial identification, eligibility criteria were applied and 39 reports were included in the meta-analysis along with our data. CDI-patients were considered as the control group. b) World map showing the countries from which the studies included in the meta-analysis come (light gray). The circles contain the number of studies selected per country.

Regarding the rest of the blood parameters evaluated, while a significant decrease in basophils count was observed **(Fig. 3 h)**, the eosinophil count was not affected by the presence of *C. difficile* **(Fig. 3 i)**. To the best of our knowledge, to date there are no reports on the role of basophils in CDI. Nevertheless, the potential role of CCL-5, a basophilic recruiter chemokine, has been highlighted in CDI ^87,88^

No differences were evident for creatinine and albumin levels **(Fig. 3 j and k)**, but an elevated urea concentration stood out in patients infected with *C. difficile* **(Fig. 3 m)**. Moreover, the BUN (Blood Urea Nitrogen)/Creatinine ratio was significantly elevated in CDI patients **(Fig. 3 n)**. Elevated BUN ratios have been associated with complications of CDI ^89^ as well as with higher mortality rates ^90^ and high urea levels were also proposed as a risk factor for severe CDI ^91^.

### Meta-analysis

We finally performed a meta-analysis to obtain more robust and reliable conclusions about the CDI associated parameters evaluated in our cohort. We defined the selection criteria (**Fig. 4 a**) and carried out a systematic review that allowed us to select 40 independent case/control type studies (**Table 2**) from the countries shown in light grey in **Fig. 4 b.**

**Table 2.**
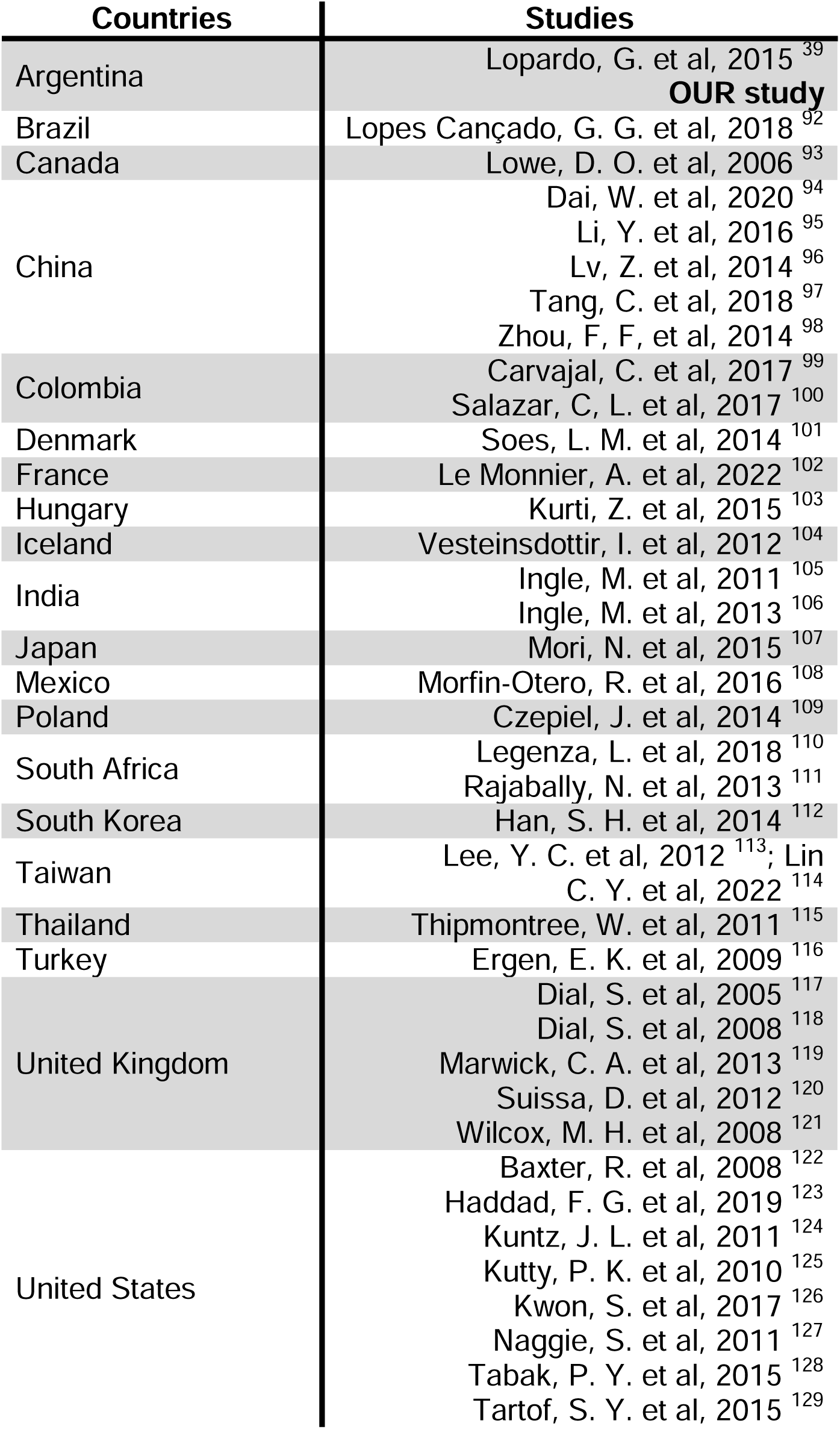
Countries and studies included in the meta-analysis.

The effects for each of the variables evaluated in the meta-analysis are summarized in **Table 3**.

**Table 3.** Summary of risk predictors from the meta-analysis.

| RISK PREDICTOR | OR |
| --- | --- |
| Age | 1,19 ** |
| Biological sex | 0,89 * |
| Prior ATB consumption | 3,02 *** |
| Prior PPI consumption | 1,29 ns |
| Prior hospitalization | 2,45 *** |
| Prior CDI | 1,76 ns |
| WBC ($10^9$ cel/mm<sup>3</sup>) | 1,35 * |
| Platelets ($10^9$ cel/ mm<sup>3</sup>) | 1,54 ns |
| Comorbidities |  |
| Diabetes *mellitus* | 1,26 ns |
| Heart disease | 1,41 * |
| Kidney disease | 2,23 *** |
| HIV | 1,00 ns |

The dataset included 11,596 individuals with CDI and 536,467 matched controls **(Table 4)**. Overall, the distribution of biological sex was 40.06% male vs. 59.94% female, being 44.33% vs. 55.67% in the CDI+ population and 39.98% vs. 60.02% in the control group. The demographic parameters Age (OR= 1.19; 95% CI, 1.06 to 1.35) and Biological Sex (OR= 0.89; 95% CI, 0.81 to 0.99) obtained significant models, which implies that, globally, advanced age and female biological sex are associated with a higher risk of CDI **(Fig. 5 a and b).**

**Figure 5.**
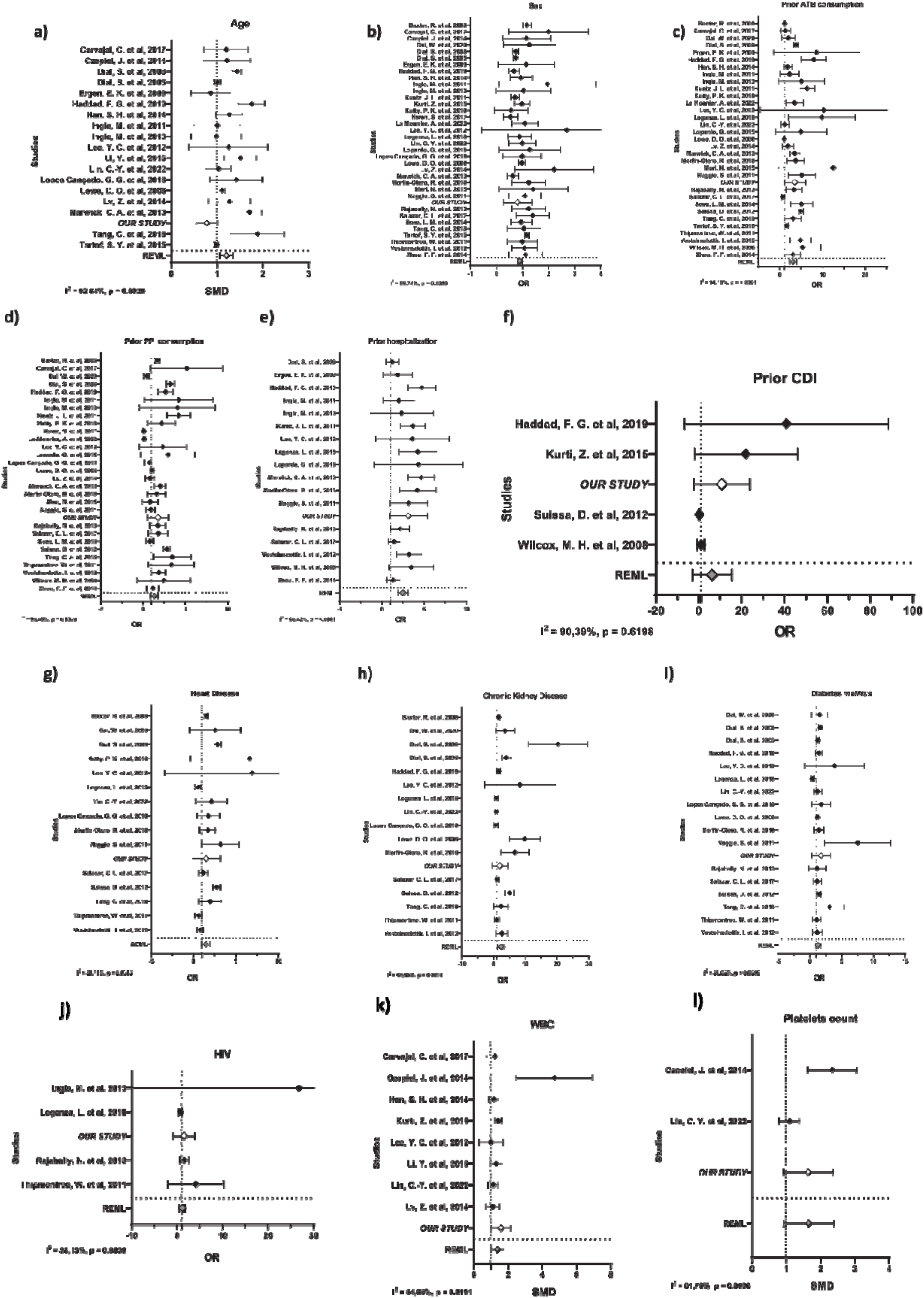
Forest plots of risk predictors. Forest plots representative of each variable evaluated in the meta-analysis. a) Age, b) biological sex, c) previous ATB consumption, d) previous PPI consumption, e) prior hospitalization, f) prior CDI, g) heart disease, h) chronic kidney disease, i) diabetes *mellitus*, j) HIV, k) white blood cells (WBC) count (cells/mm^3^), l) platelets count (cells/mm^3^). A REML (Random Effect Maximum Likelihood, gray diamond) random effects model was applied. Models with p<0.05 (OR/SMD ± CI values less or greater than 1) were considered as potential risk predictors for CDI. Black diamonds represent the means of each of the variables in each study. Bars indicate the lower and upper confidence extremes. Our study (white diamond) is mentioned as *OUR STUDY*. OR= odds ratio. SMD= standard media deviation. CI= confidence interval

**Table 4.**
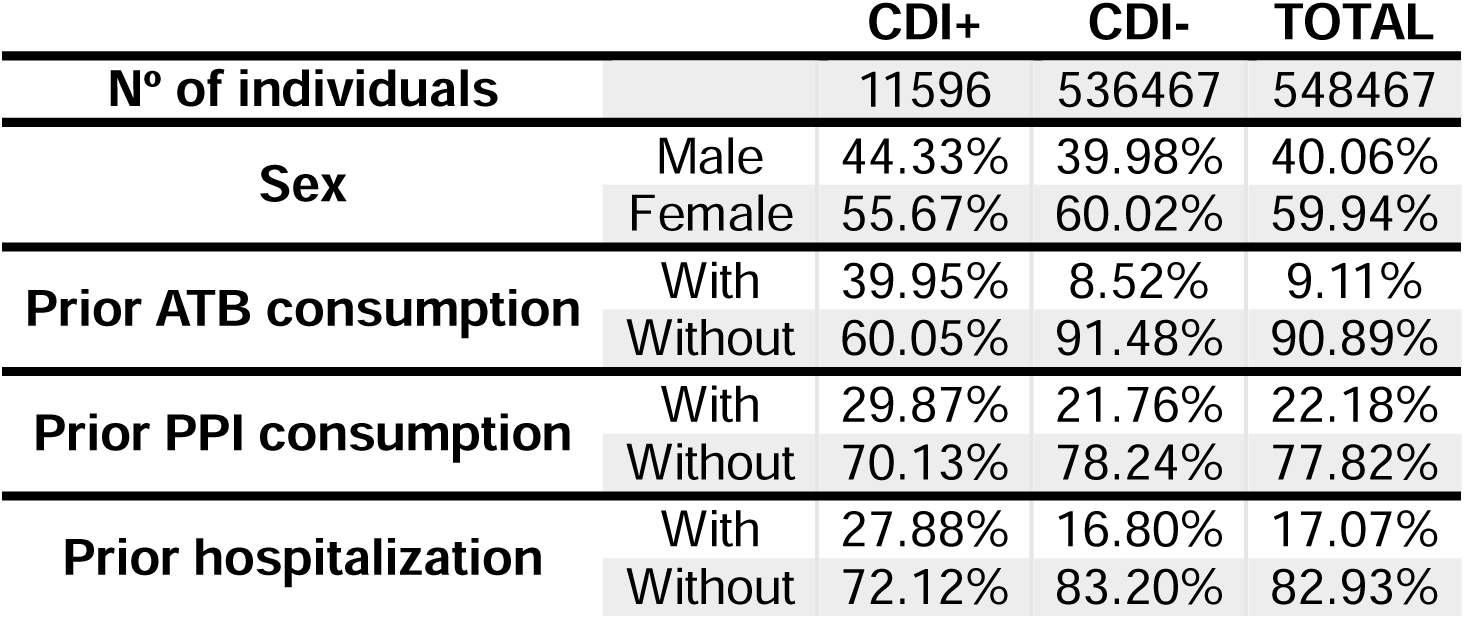
Meta-analysis dataset.

9.11% of the included individuals were previously exposed to antibiotics and 22.18% to PPI. Both antibiotic (CDI+ 39.95% vs. CDI-8.52%) and PPI (CDI+ 29.87% vs. CDI-21.76%) consumption was higher among those patients infected with *C. difficile* **(Table 4)**. In this analysis, which ignored antibiotic subclasses, we found that the pooled impact of any antibiotic exposure (OR= 3.02; 95% CI, 2.32 to 3.94) increased the risk of CDI by a multiple of 3 **(Fig. 5 c)**. A longer duration of antibiotic therapy, as well as a greater number of antibiotics administered increase the probability of CDI ^70^. The risk of acquiring CDI could be 8 to 10 times higher during antibiotic therapy, even three months after its completion, with the first month being the one with the highest risk ^130^.

Regarding PPIs as risk factors for CDI, there is an arduous discussion with some reports showing around 40-71.4% of hospitalized patients received PPI therapy during hospitalization ^131–133^ and others supporting the idea that PPIs could trigger long-term adverse effects trough changes on the microbiota composition ^134,135^. In our meta-analysis, there was no evidence of the impact of PPI on CDI risk **(Fig. 5 d)**.

Previous hospitalization emerged as the second most influential predictor for CDI risk (OR= 2.46; 95% CI, 1.90 to 3.17) **(Fig. 5 e)**. Among the CDI+ patients included in the meta-analyses, 27.88% had been hospitalized prior to diagnosis **(Table 4)** while only 16.80% in the control group.

Previous episodes of CDI are widely reported as a risk factor for subsequent cases and/or recurrences in patients. However, in this meta-analysis previous CDI was not a predictor of risk **(Fig. 5 f)** which could be attributed to the fact that only 4 of the 40 included studies provided this type of data ^56,103,120,123^. Actually, when evaluating the Forest Plot for this variable in detail, the studies show the same trend as our epidemiological study when considering previous CDI as a risk factor.

When evaluating comorbidities, both heart diseases (OR= 1.41; 95% CI, 1.02 to 1.94) and chronic kidney disease (OR= 2.23; 95% CI, 1.38 to 3.61) exhibited models with significant effects **(Fig. 5 g and h)**, indicating that these pathologies increase the risk of CDI. The rest of the tested comorbidities were not associated with a higher risk of infection **(Fig. 5 i and j)**. The presence of comorbidities has been widely reported as a condition that facilitates colonization and infection by *C. difficile* ^136^. A previous meta-analysis found IBD, diabetes, leukemia or lymphoma, kidney failure and solid cancer as the CDI risk-related comorbidities ^137^.

Finally, blood parameters were analyzed. Increased leukocytes count was also a potential predictor of CDI according to our comparative analysis (OR= 1.36; 95% CI, 1.05 to 1.75) **(Fig. 5 k)**. Although the number of platelets did not show a statistically significant effect **(Fig. 5 l)**, it is important to note that only 2 studies apart from ours evaluated this parameter.

## Conclusions

We have defined risk factors associated with CDI and detected modulations in different blood parameters in our study cohort in Argentina. We have also explored the relevance of our findings at a global level by a systematic review and meta-analysis. Our results emphasize the need to detect *C. difficile* as a causal agent of infectious diarrhea in a country where testing is not standardized or routinely performed in the health institutions. Our report provides valuable insights that could contribute to a more efficient surveillance of CDI, diagnosis and follow-up of patients.

## Author contributions

Conceptualization: all authors

Formal analysis: AMB, NDM, JC, SP, LSM and REHDP.

Funding acquisition: AMB, SP, REHDP and VP.

Investigation: AMB, NDM, SP, REHDP and VP.

Methodology: all authors.

Software: NDM and REHDP.

Supervision: REHDP and VP.

Writing: AMB wrote the original draft. All authors contributed to the review & editing of this manuscript.

## Data Availability

All data produced in the present study are available upon reasonable request to the authors.

## Acknowledgements

We thank all the patients who voluntarily participated of this study. We also thank Natalia Menite, Lucia Romano and Gastón Villafañe for their technical assistance. We acknowledge the laboratory personnel, medical staff and biochemists who have collaborated in the collection of samples and data during the study period of this report. Finally, we thank Flaticon for the icons used in the graphical abstract of this work.

