## Supplementary figures legends for "Unveiling the threat: Characterization of *Clostridioides difficile* Infection in the Northwest Region of Buenos Aires between 2019-2023 and Associated Risk Factors redefined through a Meta-Analysis"

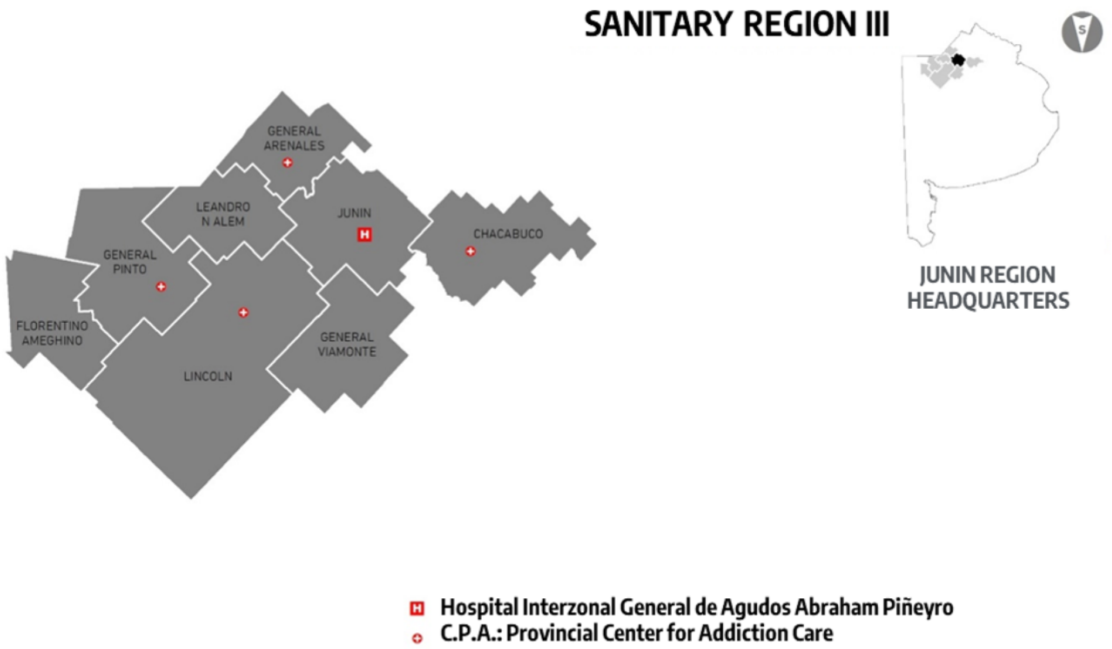


**Supplementary Figure 1.** Sanitary Region III of Buenos Aires, Argentina.

Sanitary region from the northwest of Buenos Aires province (Argentina) comprised of the municipalities of: Chacabuco, F. Ameghino, General Arenales, General Pinto, General Viamonte, Junín, Leandro N. Alem and Lincoln.

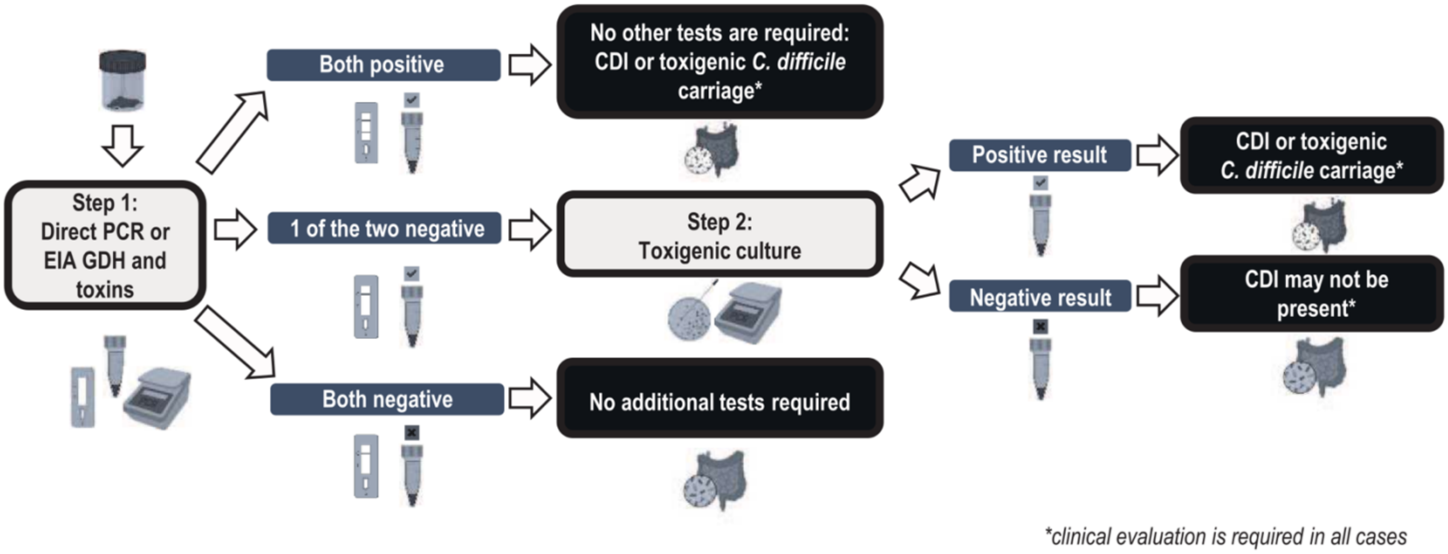


**Supplementary Figure 2.** Diagnostic algorithm to determine *C. difficile* presence in stool samples.

A cohort of 249 patients with gastrointestinal symptoms and diarrhea was evaluated. The presence of *C. difficile* within stool samples was ascertained by an algorithm that includes 3 tests (EIA, PCR, and toxigenic culture), accompanied by an exhaustive analysis of the patients' medical records. This algorithm was designed based on Crobach et al. recommendations [1].

GDH, glutamate dehydrogenase from *C. difficile*.

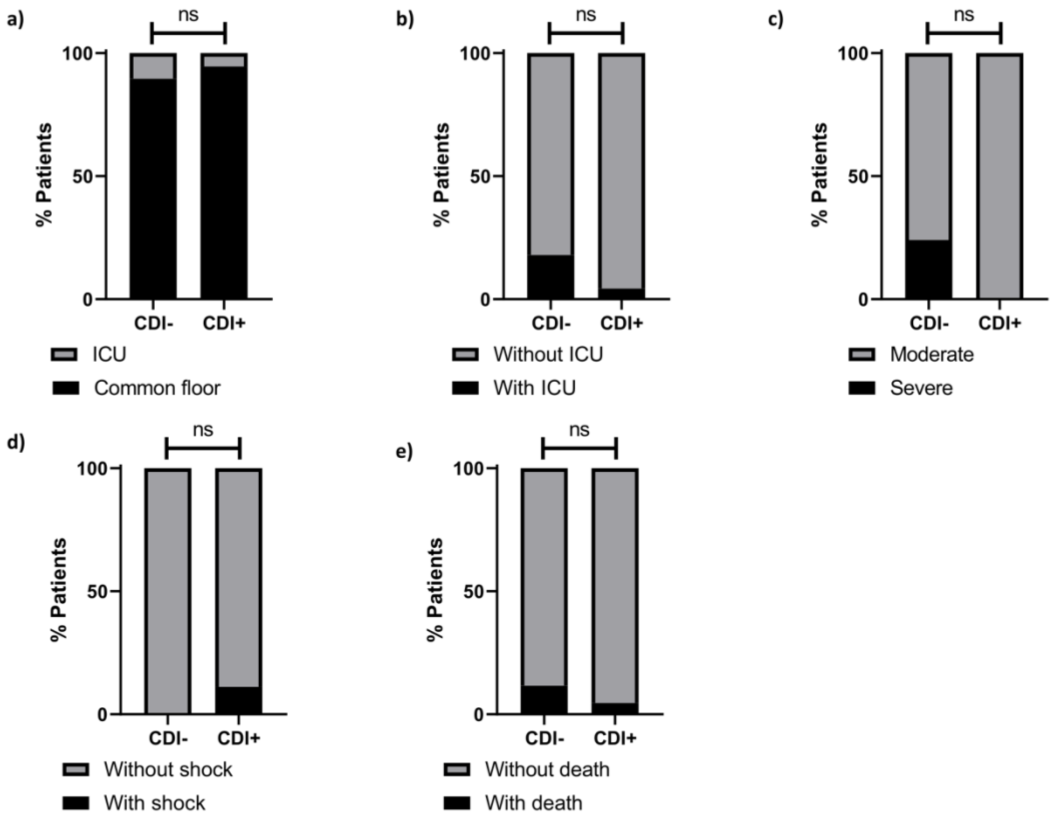


**Supplementary Figure 3.** Clinical data from CDI+ and CDI- patients.

After classifying the patients into CDI+ or CDI- populations using the diagnostic algorithm, data from clinical records was evaluated. a) Type of hospitalization room b) requirement of ICU, c) diarrhea classification d) presence of shock, e) death.

ICU= Intensive Care Unit.

a, b, c, d, e) Fisher's exact test. ns= non-significant
